# Ontology-Based SEA-CDM Integration for Vaccine Clinical Trials: Query, Similarity, and Design Recommendation

**DOI:** 10.64898/2026.11.11.26367833

**Authors:** Jiadong Zhu, Feng-Yu (Leo) Yeh, Jie Zheng, Yongqun (Oliver) He

## Abstract

ClinicalTrials.gov provides valuable information for vaccine research, but fragmented data structures and inconsistent vaccine naming complicate systematic cross-trial analysis. The Study-Experiment-Assay Common Data Model (SEA-CDM) provides a standardized structure for biomedical study data, while biomedical ontologies supply semantic relationships among concepts. We extracted 9,147 vaccine-related study records from the Aggregate Analysis of ClinicalTrials.gov (AACT) database, mapped their records into SEA-CDM, and linked vaccine products and reported adverse events (AEs) to the Vaccine Ontology (VO) and Ontology of Adverse Events (OAE), respectively. Building on the existing Ontology-based SEA Network (OSEAN), we organized the integrated data into relational tables and a Neo4j knowledge graph while retaining links to original registrations. We evaluated three applications to assess semantic trial retrieval, context-aware AE ranking and trial similarity, and trial design recommendation. First, VO-based query expansion retrieved 41 candidate RNA vaccine trials compared with 16 using exact VO identifier matching. Second, a context-aware AE ranking model incorporating SEA-CDM features and VO-derived vaccine families improved recall@10 from 0.317 to 0.383 over a frequency-based baseline. TransE and CompGCN embeddings achieved Hits@10 of 0.950 and 0.946, respectively, for retrieving trials sharing the same VO identifier within disease-specific candidate sets. Third, an ontology-based clinical trial design recommendation model was developed using several machine learning methods under leave-one-Material-out evaluation across 23 vaccine classes, although performance varied across classes. These findings demonstrate how structural and semantic integration can support multiple clinical trial analyses using vaccine studies as a case study. This framework provides a foundation for extending ontology-supported data integration and analysis to other clinical research domains.

## 1. Introduction

ClinicalTrials.gov provides valuable information for vaccine clinical trial research, including vaccine products, study designs, participant characteristics, and reported adverse events (AEs). Researchers and vaccine developers often need to identify relevant historical trials, compare reported AEs across similar studies, and examine previously registered trial designs. The Aggregate Analysis of ClinicalTrials.gov (AACT) database makes these data available in a relational format [1,2]. However, relevant information is distributed across multiple tables, and vaccine products are often recorded as free text, making systematic data integration and cross-trial analysis challenging. Without standardized vaccine identifiers and explicit biological relationships, identifying similar vaccines often relies on text matching or computational similarity measures that may not capture their underlying biological connections.

Biomedical ontologies provide standardized concepts and relationships that support semantic data integration and analysis, following principles promoted by initiatives such as the OBO Foundry [3] and FAIR data management practices [4]. The Vaccine Ontology (VO) systematically represents vaccine products, vaccine types, and their biological relationships, covering licensed vaccines, investigational vaccines, and vaccines evaluated in animal models [5–7]. For example, an RNA vaccine and a more specific COVID-19 RNA vaccine may have different VO identifiers, but their hierarchical relationship allows related trials to be retrieved together. VO is also linked to the Vaccine Investigation and Online Information Network (VIOLIN), a curated vaccine knowledge base that provides additional product information and stable vaccine identifiers [8,9]. However, mapping free-text vaccine names from clinical trial records to VO remains challenging and has motivated the development of specialized natural language processing approaches [10]. Similarly, the Ontology of Adverse Events (OAE) provides standardized representations and hierarchical relationships for adverse events reported following medical interventions, including vaccination [11–13]. Together, VO and OAE provide a semantic foundation for organizing vaccine products and reported AEs.

While biomedical ontologies support semantic standardization, they do not by themselves provide a unified structure for organizing all components of clinical trial records. Existing common data models, such as the OMOP Common Data Model developed within the Observational Health Data Sciences and Informatics (OHDSI) community, primarily support the standardization of observational healthcare data [14,15]. In contrast, ClinicalTrials.gov contains heterogeneous registry information, including study protocols, interventions, eligibility criteria, and reported results, which presents additional challenges for secondary analysis [16]. Previous studies have demonstrated the value of knowledge graphs and embedding-based approaches for clinical trial retrieval and recommendation [17,18]. However, previous approaches have not typically integrated vaccine-specific ontology relationships with a standardized clinical trial data model to support multiple downstream applications within a unified framework.

The ontology-supported Study–Experiment–Assay Common Data Model (SEA-CDM) is a newly proposed cross-domain common data model that provides a standardized schema for integrating biomedical study evidence across domains [19]. SEA-CDM consists of 10 core tables and three accessory tables, providing a relatively compact schema for biomedical data integration. The advantage of SEA-CDM has been verified using vaccine transcriptomics data [19]. AACT organizes ClinicalTrials.gov records across 51 tables, requiring joins across multiple tables for certain cross-trial analyses, which makes query and analysis difficult. For example, identifying adverse events reported in trials involving related RNA vaccines may require linking intervention, trial arm, and adverse event records across multiple AACT tables, while also accounting for inconsistent vaccine names. In the proposed framework, these records are organized into linked SEA-CDM entities and grounded to VO and OAE concepts, allowing related vaccine trials and their reported adverse events to be retrieved through ontology-based queries rather than exact text matching alone. We hypothesized that integrating vaccine clinical trial data with SEA-CDM and biomedical ontologies could facilitate systematic data integration, semantic querying, and downstream machine learning analysis.

To address the above hypothesis, we developed an automated pipeline to extract vaccine-related clinical trial records from AACT, map them into SEA-CDM, and link vaccine products and reported AEs to VO and OAE, respectively. Building on the previously established Ontology-based SEA Network (OSEAN) framework [19], we organized the integrated vaccine clinical trial data into relational and knowledge graph representations. We evaluated three complementary applications: (i) query—using VO expansion to retrieve related vaccine trials and examine OAE-grounded AE information; (ii) similarity and prediction—combining SEA-CDM context and VO-derived information for AE ranking, together with knowledge graph embeddings for trial similarity analysis; (iii) clinical trial design recommendation—using ontology-based similarity to identify relevant historical trials and suggest registered design characteristics. Together, these applications demonstrate how a standardized clinical data model and biomedical ontologies can be integrated to support multiple types of vaccine clinical trial analysis.

## 2. Materials and Methods

**Figure 1** illustrates the overall workflow of this study. First, vaccine-related clinical trial records were extracted from the AACT relational database [1,2]. The extracted records were then converted into SEA-CDM to provide a standardized representation of clinical trial information. Vaccine products and platforms were mapped to the Vaccine Ontology (VO), with VIOLIN product identifiers used when available, while reported adverse events (AEs) were mapped to the Ontology of Adverse Events (OAE). ClinicalTrials.gov identifiers (NCT IDs) were retained to link the integrated data to their original trial records. The resulting database and knowledge graph supported three downstream applications: ontology-based query, similarity and AE ranking, and clinical trial design recommendation. All analyses were based on the SEA-CDM AACT vaccine remapped v1.4 dataset, containing 9,147 studies from the AACT snapshot dated April 12, 2026, and the VO release dated June 12, 2026.

**Figure 1.**
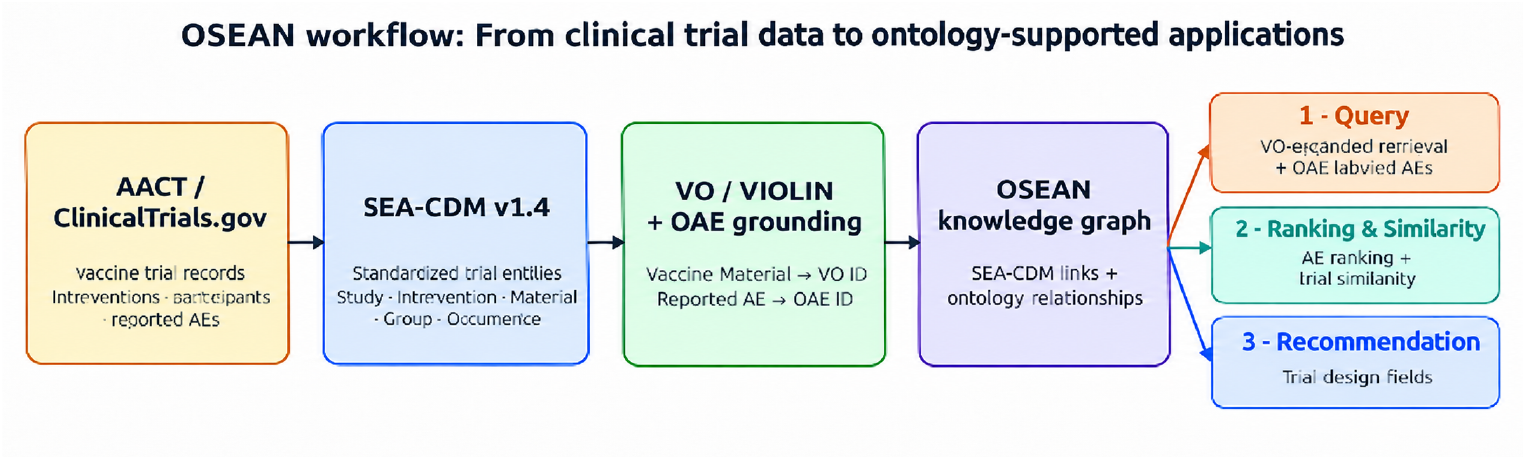
General project workflow.

### 2.1 Data conversion using ontology-based SEA-CDM

#### Vaccine trial selection and inclusion criteria

Vaccine-related clinical trial records were identified from the local AACT pipe-delimited export using a predefined keyword-based filtering procedure implemented in the VaxCT preprocessing pipeline. Studies were included if at least one case-insensitive, whole-word match to a vaccine- or immunization-related term was found in any of three AACT fields: `studies.brief_title`, `interventions.name`, or `keywords.name`. The term list included general vaccine and immunization terms (e.g., vaccine, vaccination, immunization, and immunisation), as well as selected product-, platform-, and disease-related terms (e.g., mRNA, pneumococcal, HPV, and BCG). The complete keyword list is provided in **Table S1**.

Matching was performed using word-boundary regular expressions to reduce unintended substring matches. The final cohort was defined as the union of NCT identifiers retrieved from the three fields. All AACT tables required for subsequent SEA-CDM mapping were restricted to the selected NCT identifiers.

No additional requirement for an identified investigational vaccine product was imposed during cohort selection. Therefore, some included studies may lack a specific product-level VO mapping. The final NCT cohort and the filtering procedure were retained to support reproducibility.

#### SEA-CDM construction

SEA-CDM provides a standardized structure for organizing biomedical study information into defined entities and relationships, including Study, Experiment, Intervention, Material, Subject, Group, Occurrence, and Result [19]. SEA-CDM adopts a study-centered structure that explicitly organizes interventions, participant groups, and reported outcomes. This structure is particularly useful for representing ClinicalTrials.gov records, where trial design information and reported results must be integrated within a common framework.

We adapted this structure to represent the selected vaccine-related clinical trial records from AACT. Each included ClinicalTrials.gov registration was represented as a Study identified by its NCT ID, with the associated registry context organized under Experiment. Identifiable investigational vaccine products were represented as SEA-CDM Material entities, with each mapped vaccine product linked to a primary VO identifier. Vaccination procedures were represented as Intervention entities. Eligibility and demographic information were organized under Subject, trial arms under Group, and reported adverse events under Occurrence. Study design and protocol information was also retained as additional attributes for downstream analysis. Assay and Sample entities remained in the schema for compatibility with the original SEA-CDM model, although these data were less frequently available in ClinicalTrials.gov records and were not required for the primary analyses (**Figure 2**).

**Figure 2.**
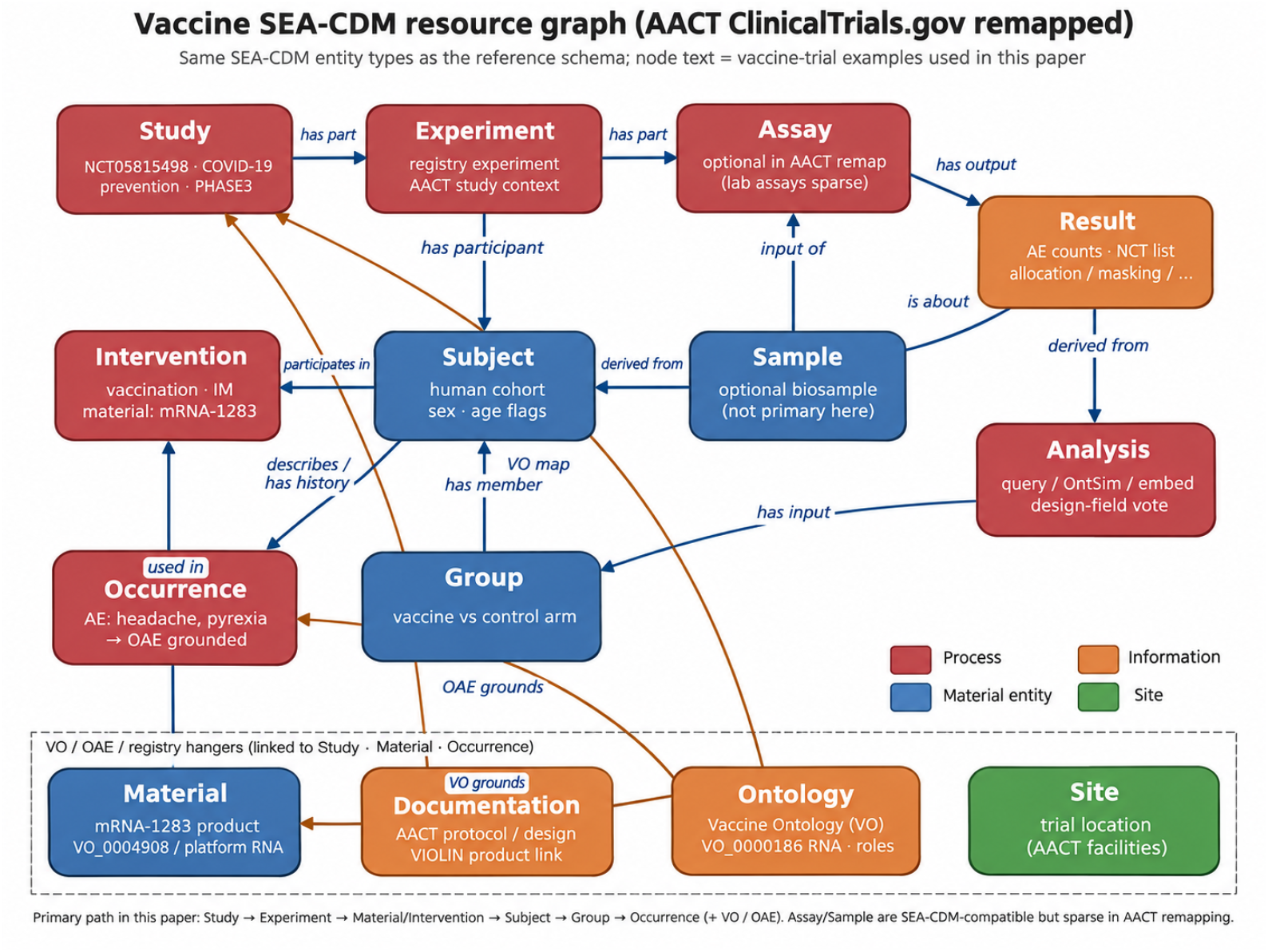
Representation of a vaccine clinical trial using the SEA-CDM schema and its links to vaccine and adverse event ontologies.

The integrated records were represented in relational SEA-CDM tables and an NCT-linked knowledge graph implemented using Neo4j [20]. The relational representation supports structured SQL queries, while the knowledge graph enables the exploration of relationships among trial entities and ontology concepts. Additional study characteristics, including disease, phase, study purpose, participant demographics, administration route, dosage, and VO mappings, were retained for downstream analyses. Feature sources are summarized in **Table S2**, while resource counts and OAE coverage are provided in Section S3.

#### VO/OAE grounding and ontology graph integration

To provide standardized semantic representations, vaccine products and reported AEs were linked to their corresponding ontology concepts. VO organizes vaccine products and types through hierarchical and role-based relationships, including `subClassOf` and `has_role` [5,6]. VIOLIN provides additional curated vaccine product identifiers, such as `VIOLIN_5784` for Comirnaty, which can support the mapping of registry vaccine names to VO concepts [8].

Similarly, OAE provides standardized classes and hierarchical relationships for adverse events [11,12]. Reported AE terms were mapped to OAE identifiers using label and synonym matching, supplemented by manually defined mappings for selected ambiguous terms, such as pyrexia to fever AE. In our pipeline, each identified investigational vaccine product was assigned a primary VO identifier and, where applicable, a broader vaccine platform identifier. Details of the vaccine-to-VO mapping audit and subsequent corrections are provided in Section S8.

Importantly, the integrated knowledge graph incorporated not only the mapped VO and OAE identifiers but also their surrounding ontology relationships. Vaccine products were linked to VO classes, while reported AE terms were linked to OAE classes through SEA-CDM Occurrence records. The ontology hierarchy was preserved through relations such as `subClassOf`, allowing related vaccine categories and AE concepts to be explored within the same knowledge graph. This integrated representation supported the three downstream applications.

### 2.2 Use case 1 — Query

Use Case 1 focuses on retrieving vaccine clinical trials and their reported adverse events (AEs) using the integrated SEA-CDM data. We implemented two query approaches: exact VO identifier matching (Layer A) and ontology-based expansion (Layer B).

#### Layer A (Exact VO identifier matching)

In Layer A, trials are retrieved by matching the primary VO identifier assigned to an investigational vaccine product (`arch_vo` = *v*). Related SEA-CDM entities, including Intervention, Subject, Group, and Occurrence, are then used to examine reported AEs and filter trials by characteristics such as administration route, age, sex, or trial arm. When an AE term is successfully mapped to OAE, its standardized OAE identifier and label are displayed alongside the number of matching trials.

#### Layer B (Ontology-based expansion)

Unlike exact matching, Layer B considers related vaccine classes connected through VO relationships. For example, a query for RNA vaccines can also retrieve trials assigned to more specific classes, such as COVID-19 RNA vaccines. Instead of requiring `arch_vo`= *v*, the query expands the selected VO class to include related ontology concepts. Let *N* ^↑^(*v*) denote ancestor classes and role-related peers, and *D*^↓^(*v*) denote descendant classes up to depth three, with sibling expansion disabled by default:

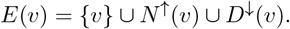

Trials are included when their VO identifiers belong to the expanded set *E*(*v*). Exact VO identifier matching (`arch_vo` = *v*) serves as the comparison baseline.

Both approaches support five query patterns: forward, reverse, compare, drill-down, and conditional queries. For AE ranking, the frequency of an AE term *a* is calculated as:

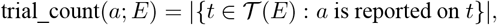

where *T* (*E*) represents the set of trials satisfying the VO identifier filter and any additional Intervention, Subject, or Group constraints. These counts represent the number of matching trials reporting each AE, rather than predicted AE incidence rates. Representative results for exact VO identifier queries and ontology-expanded queries are provided in **Table S6** and **Table 1**, respectively.

**Table 1.**
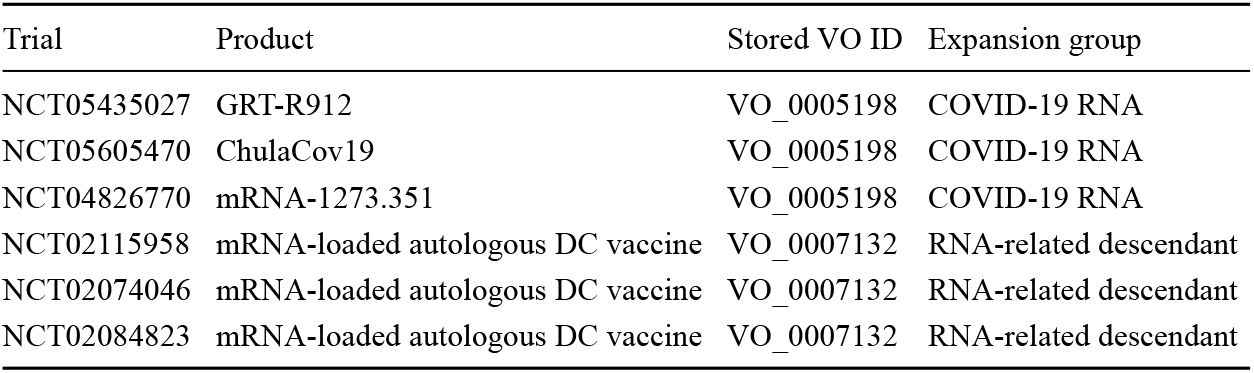
Representative trials retrieved through VO expansion but not Exact VO identifier matching for RNA vaccines (VO_0000186).

### 2.3 Use case 2 — Similarity and prediction

Use Case 2 evaluates two complementary tasks using the integrated OSEAN data: context-aware AE ranking and knowledge graph-based trial similarity.

#### AE ranking (primary task)

The goal of AE ranking is to identify which adverse events are most likely to be reported in a given trial context. Each example corresponds to an NCT *×* Group-arm context *x*, associated with a set of reported AE terms *Y*_*x*_. Contexts were restricted to NCT × vaccine/control arms with at least one reported AE in the SEA-CDM Occurrence records; from these arms we used the 80 most frequent AE terms that each appeared in at least 25 contexts, yielding 2,571 contexts. The data were divided into training (80%) and testing (20%) sets by study, ensuring that all arms from the same trial were assigned to the same split.

We compared two models. Model A ranks AEs according to their overall frequency in the training data, producing the same ranking for all trial contexts. Model B generates context-specific AE rankings using vaccine family and SEA-CDM study characteristics. These models are defined as:

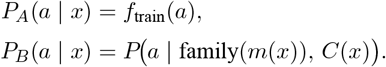

where *m*(*x*) represents the vaccine’s primary VO identifier and *C*(*x*) includes administration route, dosage, sex, age, trial arm role, and disease indication. The vaccine family is derived from VO relationships, including `subClassOf` and `has_role`, which connect specific vaccine products to broader platform categories. Model B uses one-vs-rest logistic regression [21].

Reported and predicted AE terms were linked to OAE identifiers when mappings were available (78 of 80 terms). These mappings support interpretation of the results but are not used as prediction features. Model performance was evaluated using recall@10:

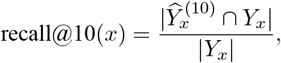

where *Y*_*x*_ represents the ten highest-ranked predicted AE terms. This metric measures the proportion of reported AEs recovered among the top ten predictions.

#### Trial similarity (secondary task)

To identify related vaccine trials, we generated study embeddings from the SEA-CDM and VO knowledge graph using TransE [22] and CompGCN [23]. Trial similarity was calculated using cosine similarity between study embeddings. For each query trial, candidate trials were ranked according to their embedding similarity, with trials sharing the same VO identifier considered relevant neighbors.

After joining the analysis cohort to VO-mapped vaccine products, 556 studies were available for embedding-based similarity. Evaluation used disease-specific candidate sets (other studies with the same disease indication): queries without a usable same-disease candidate set were excluded, leaving 541 queries. We reported Hits@1, Hits@10, and Mean Reciprocal Rank (MRR). Hits@K measures the proportion of queries for which at least one relevant trial appears among the top *K* ranked candidates, while MRR summarizes the reciprocal rank of the first relevant trial. A VO-only edge ablation is provided in Section S5. Although OAE relationships were included in the integrated knowledge graph for AE-related analysis, they were not required for these trial similarity evaluations.

### 2.4 Use case 3 — Design recommendation

We developed an ontology-based clinical trial design recommendation model that identifies similar historical trials and uses their registered designs to recommend design characteristics. The model predicts four design fields: allocation, masking, intervention model, and enrollment size category. Recommendations are based on disease, study phase, study purpose, and vaccine product and platform VO identifiers.

For each query trial, the system identifies the top 20 most similar training trials and predicts each design field through a similarity-weighted vote. The design fields serve as prediction targets rather than input features or graph edges.

#### Neighbor scoring

The similarity between a query trial *q* and a historical trial *j* is calculated as:

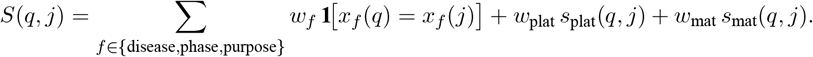

We assigned weights of 1 to disease and phase, 0.5 to study purpose, 1 to platform similarity, and 0.5 to vaccine-class similarity.

We compared exact identifier matching with ontology-based similarity. Exact matching assigns a similarity of 1 when two VO identifiers are identical and 0 otherwise:

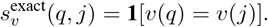

Ontology-based similarity (OntSim) instead measures the overlap between the VO neighborhoods of two vaccine or platform VO classes using Jaccard similarity:

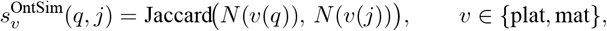

where *N* (·) represents the set of related ontology concepts obtained through `subClassOf` and `has_role` relationships. Identical identifiers receive a similarity score of 1.

Unlike exact matching, OntSim allows related vaccine classes with different VO identifiers to contribute to the recommendation. Disease, phase, and purpose matching remain the same for both approaches. The final prediction for each design field is determined by a similarity-weighted vote among the top 20 neighbors.

#### Evaluation

We used two evaluation strategies to assess performance under different scenarios: prediction for future trials and generalization to previously unseen vaccine classes.

#### Chronological evaluation (past to future)

Trials with start dates before 2022 were used for training, while trials starting in 2022 or later were used for testing (n=283). This evaluation simulates using historical trial information to recommend designs for future studies. Because many VO identifiers assigned to test trials were already represented in the training data, exact matching and ontology-based similarity could identify similar neighbors with identical VO identifiers.

#### Leave-one-Material-out (LOMO) evaluation

To assess generalization to previously unseen vaccine classes, we evaluated 23 ontology-connected vaccine classes, each defined by a primary VO identifier. In each round, all trials assigned to one vaccine class were excluded from training and used for evaluation (Section S4). Under this setting, exact matching cannot identify training trials with the same VO identifier, whereas OntSim can retrieve trials involving related vaccine classes through VO relationships. This evaluation therefore examines whether ontology relationships provide additional information when exact VO identifier matches are unavailable.

The reference labels were obtained from registered clinical trial designs. Accordingly, the recommendations reflect historical design choices rather than clinically optimal designs. Additional baseline methods and a prototype user interface are presented in Section S2 and **Figure S1**. The main results are presented in **Table 4** and **Figure 6**.

## 3. Results

The integration of vaccine clinical trial records produced a standardized SEA-CDM resource with ontology-grounded study information. From 579,828 AACT study registrations, 9,147 vaccine-related studies were identified and mapped into SEA-CDM, yielding 18,294 Experiment records, 20,001 Intervention records, 9,147 Subject records, 617,510 Occurrence records, and 1,525,757 Result records. Among 14,190 distinct registry-reported AE terms, 5,040 were mapped to OAE concepts. The resulting relational database and knowledge graph provided a shared representation for the three downstream applications.

### 3.1 Query: Ontology expansion beyond exact VO identifier matching

VO-based query expansion retrieved more RNA vaccine trials than exact VO identifier matching. Using RNA vaccines (`VO_0000186`) as an example, exact matching identified 16 trials, whereas ontology expansion retrieved 41 candidate trials, including 25 additional trials (**Figure 3**). Exact matching retrieves only trials assigned to the queried VO identifier, while ontology expansion also includes trials mapped to related vaccine classes.

**Figure 3.**
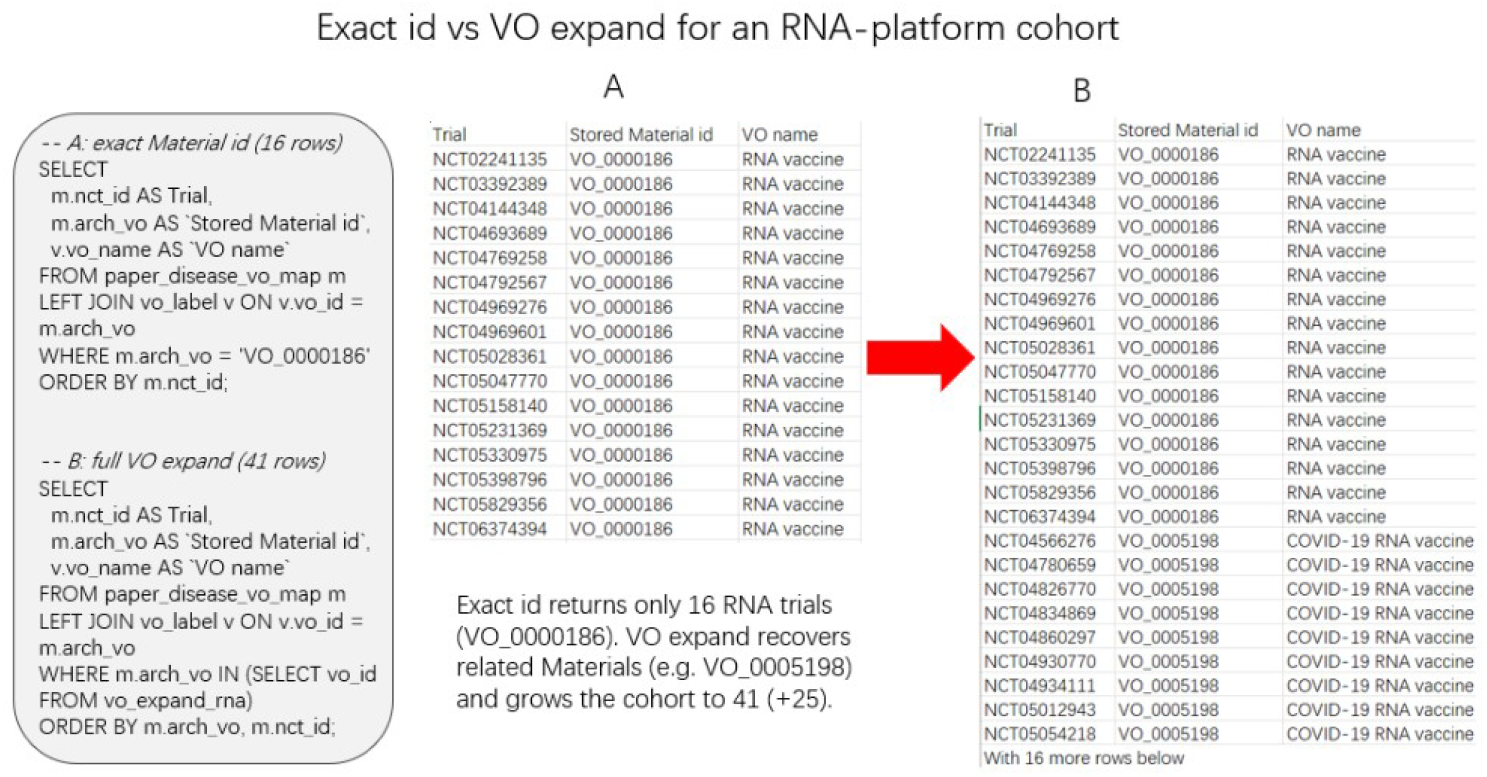
Comparison of Exact VO identifier matching and VO-based query expansion for RNA vaccines (VO_0000186). (A) Exact matching retrieves 16 trials assigned to the queried VO identifier. (B) Ontology expansion retrieves 41 candidate trials, including 25 additional trials mapped to related vaccine classes.

**Table 1** presents representative trials identified only through ontology expansion. Among the 25 additional records, 20 were mapped to the COVID-19 RNA vaccine class (`VO_0005198`), and five were mapped to the mRNA-loaded autologous dendritic cell vaccine class (`VO_0007132`). These trials were not retrieved through exact matching because their stored VO identifiers differed from the queried RNA vaccine identifier. The complete list of additional trials is provided in **Table S8**.

These results demonstrate how VO relationships can broaden clinical trial retrieval beyond exact identifier matching, allowing related vaccine classes to be included in the same query. Exact VO identifier queries can also be used to examine reported AEs for a specified vaccine class, as illustrated in **Table S6**.

### 3.2 Similarity and prediction: context-aware AE ranking

Combining SEA-CDM study characteristics with VO-derived vaccine families improved AE ranking compared with a frequency-based baseline. Model A produces the same AE ranking for all trials, whereas Model B adjusts the ranking based on vaccine family, administration route, disease indication, and other study characteristics. Model B achieved a recall@10 of 0.383, compared with 0.317 for Model A (Δ = +0.066; **Table 2**). This means that Model B recovered a larger proportion of reported AEs among its top 10 predictions for each trial context than the frequency-based baseline.

**Table 2.** Performance of AE ranking and trial similarity methods. AE ranking compares a frequency-based baseline (Model A) with the context-aware Model B. Trial similarity evaluates TransE and CompGCN using shared VO identifiers as the relevance criterion. A dash indicates that the metric is not applicable.

| Task | Setting | Hits@1 | Hits@10 | MRR | recall@10 |
|---|---|---|---|---|---|
| AE ranking | Model A (frequency baseline) | — | — | — | 0.317 |
| AE ranking | Model B (SEA-CDM + VO) | — | — | — | 0.383 |
| Trial similarity | TransE full (SEA-CDM + VO) | 0.571 | 0.950 | 0.710 | — |
| Trial similarity | CompGCN full (SEA-CDM + VO) | 0.575 | 0.946 | 0.688 | — |

Knowledge graph embeddings effectively retrieved trials sharing VO identifiers within the evaluated disease-specific candidate sets. Across 541 queries, TransE and CompGCN achieved Hits@10 values of 0.950 and 0.946, respectively, indicating that at least one trial sharing the query’s VO identifier appeared among the top 10 candidates for approximately 95% of queries (**Table 2**). TransE achieved an MRR of 0.710, compared with 0.688 for CompGCN. A VO-only edge ablation is presented in Section S5.

Model B coefficients revealed how structured SEA-CDM characteristics and VO-derived features contributed to context-specific AE rankings (**Figures 4 and 5**). VO features showed the largest aggregate coefficient magnitude, followed by study indication and intervention features (**Figure 4**). Positive coefficients indicate higher predicted scores for the corresponding AEs in a given trial context, whereas negative coefficients indicate lower scores. For example, RSV indication was positively associated with cough, rhinorrhoea, and diarrhoea, while intranasal administration showed a negative coefficient for pneumonia (**Figure 5**). These patterns illustrate how the model adjusts AE rankings according to trial characteristics rather than relying solely on overall reporting frequency.

**Figure 4.**
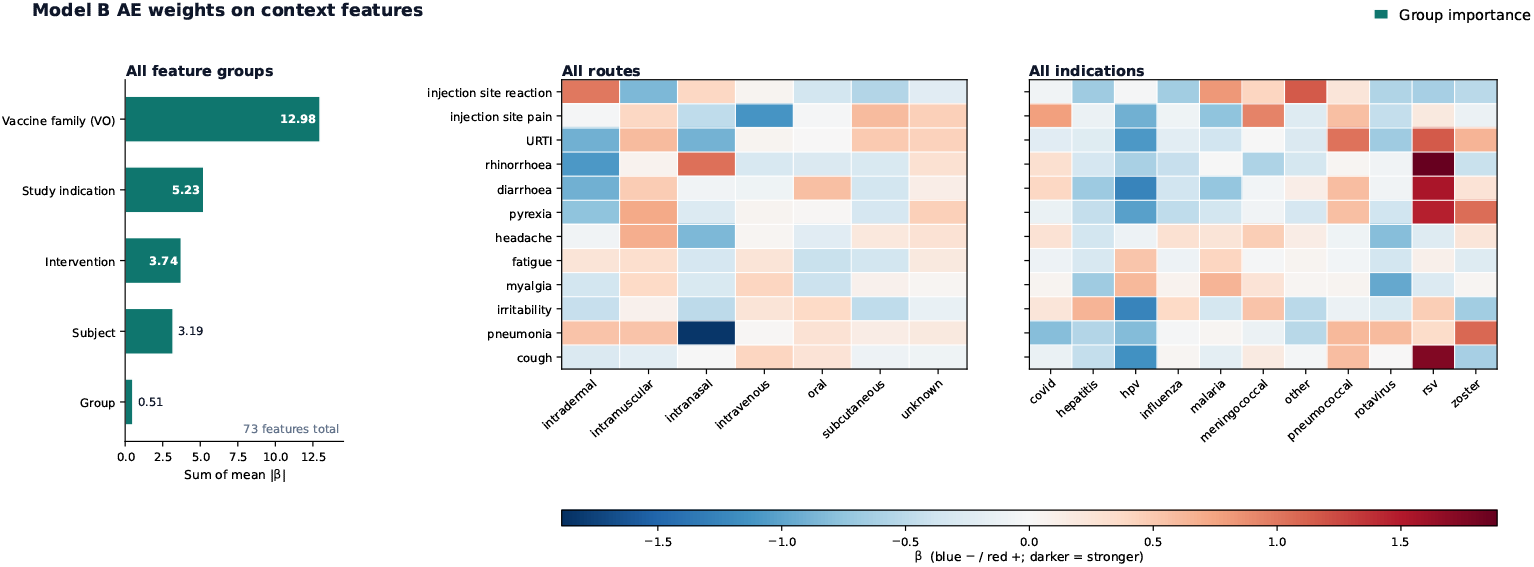
Logistic regression coefficients of Model B across SEA-CDM/VO feature groups, administration routes, and disease indications. The left panel summarizes coefficient magnitudes by feature group, while the heatmaps display coefficients for selected AEs (blue: negative; red: positive; darker colors indicate larger magnitudes).

**Figure 5.**
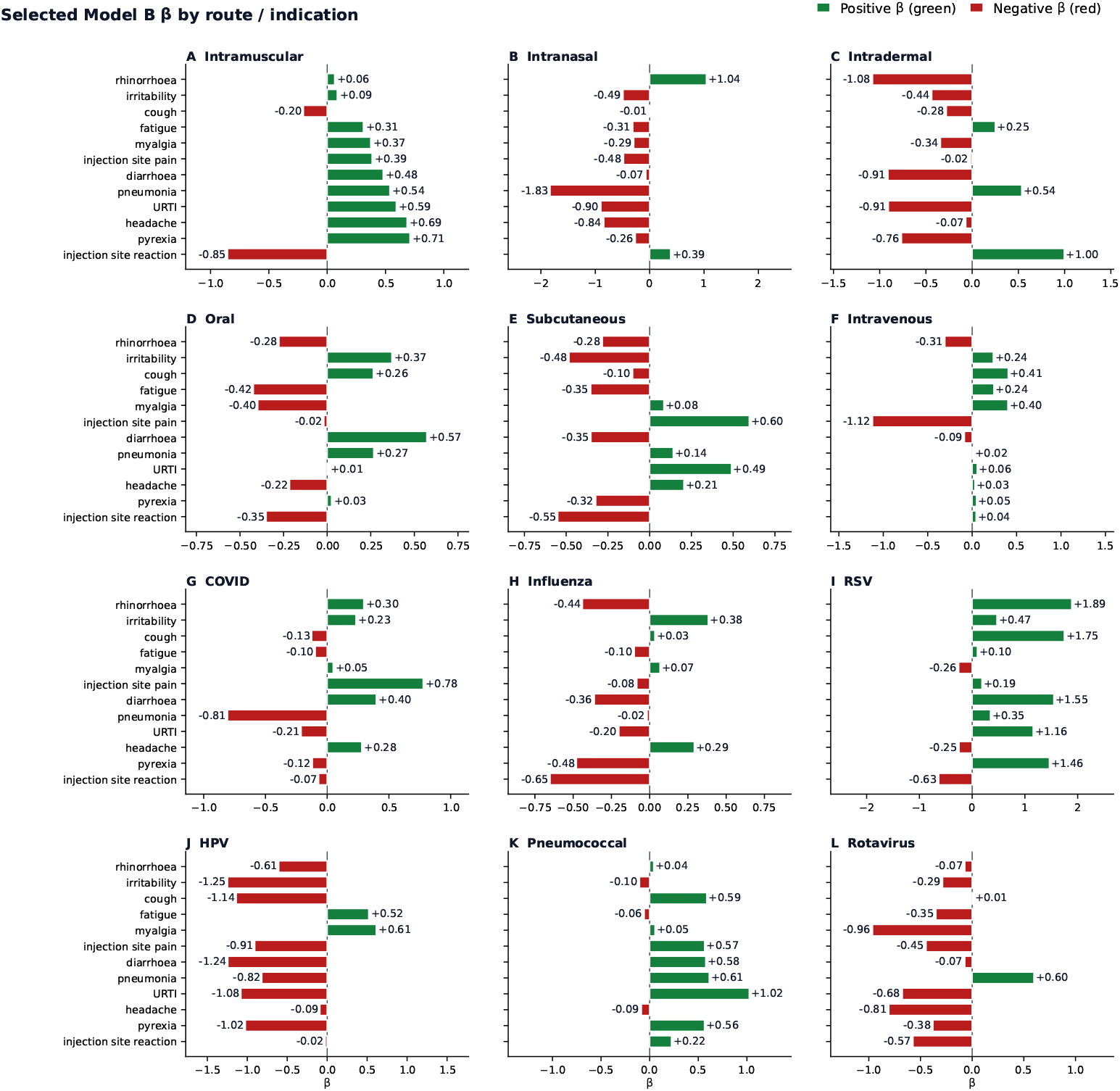
Selected logistic regression coefficients of Model B for six administration routes and six disease indications. Positive and negative coefficients are shown for the same AE terms as in Figure 4. Each panel uses an independently scaled x-axis to improve readability.

**Figure 6.**
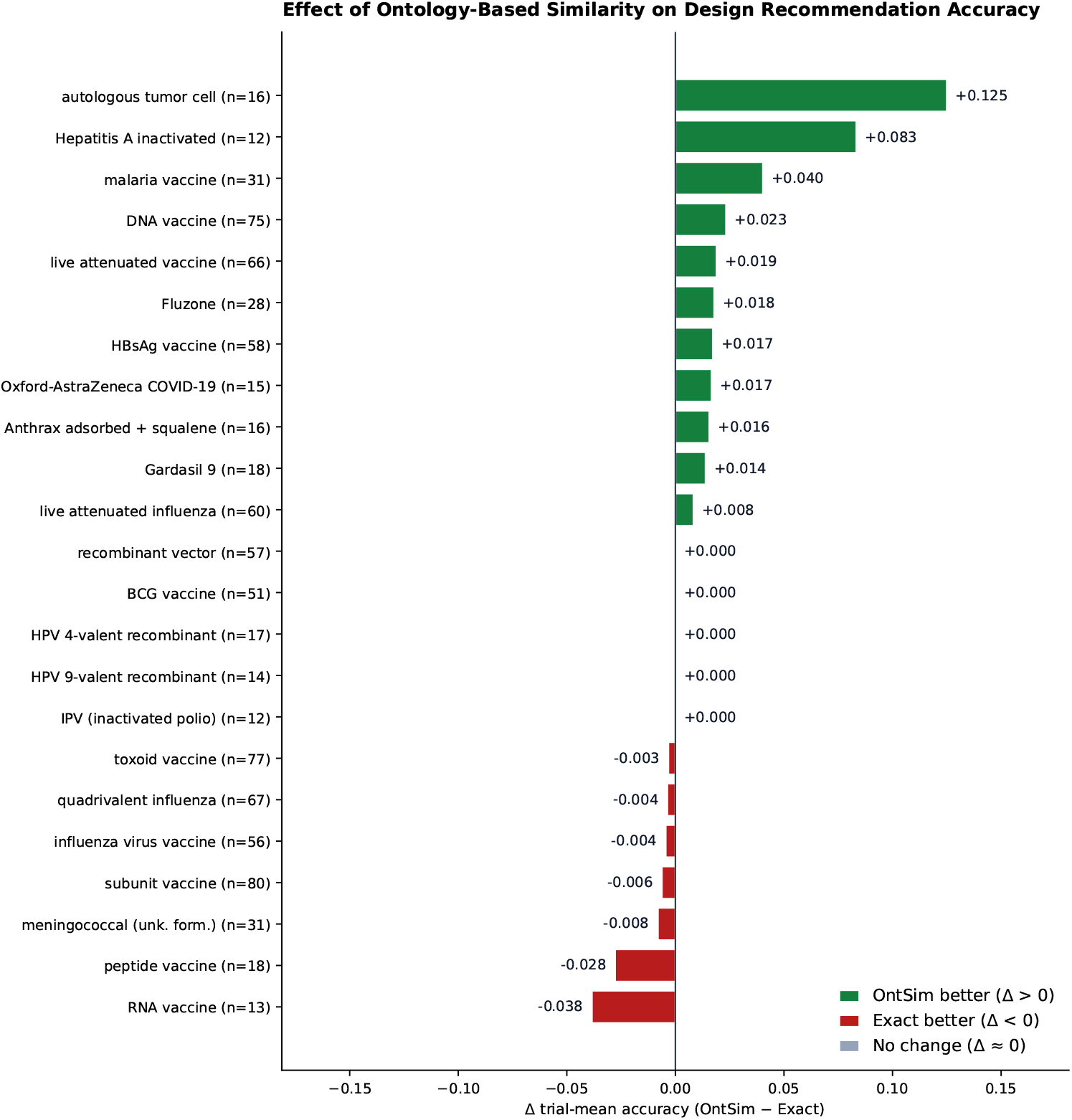
Per-class difference in trial-mean accuracy between OntSim and Exact under leave-one-Material-out evaluation across 23 vaccine classes. Positive values indicate better performance with OntSim, while negative values favor Exact matching. Overall class-equal accuracy was 0.646 for OntSim and 0.633 for Exact (Δ = +0.013).

To illustrate how trial context influences AE ranking, **Table 3** presents a Comirnaty-style query using the nucleic-acid vaccine family, intramuscular administration, COVID-19 indication, and adult participants. Compared with Model A, Model B moved upper respiratory tract infection from rank 5 to rank 2, diarrhoea from rank 10 to rank 4, and injection site pain from rank 11 to rank 6, while headache remained the highest-ranked AE. This example illustrates how incorporating vaccine family and structured trial characteristics changes the predicted AE ranking relative to overall reporting frequency. The Model B scores are used for ranking and should not be interpreted as clinical AE incidence probabilities.

**Table 3.** Example of context-aware AE ranking for a Comirnaty-style trial query. Model B ranks AEs using vaccine family and study characteristics, while Model A ranks them according to overall training-set frequency. Model B scores are relative ranking scores that order predicted AEs for the queried trial context.

| Rank | Predicted AE (Model B) | Model B score | Baseline rank (Model A) | OAE id |
|---|---|---|---|---|
| 1 | Headache | 0.921 | 1 | OAE_0000377 |
| 2 | Upper respiratory tract infection | 0.752 | 5 | OAE_0000358 |
| 3 | Fatigue | 0.558 | 2 | OAE_0000034 |
| 4 | Diarrhoea | 0.527 | 10 | OAE_0000616 |
| 5 | Myalgia | 0.517 | 3 | OAE_0000375 |
| 6 | Injection site pain | 0.515 | 11 | OAE_0006886 |

**Table 4.**
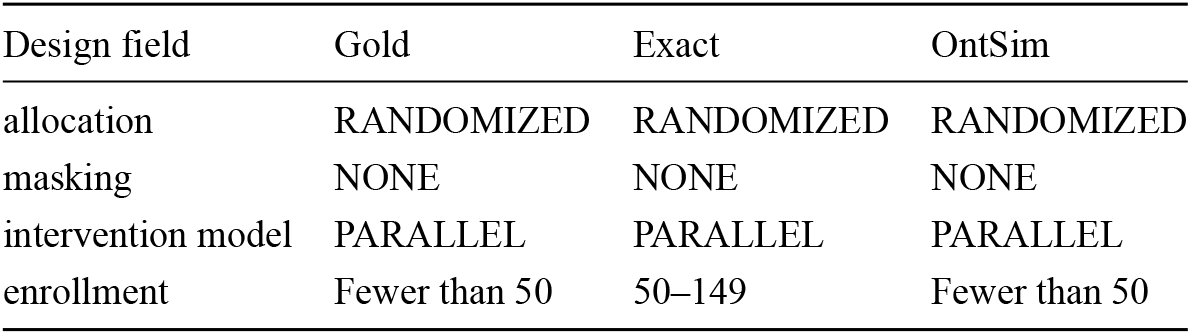
Clinical trial design recommendation for NCT00799110 under leave-one-Material-out evaluation (*k* = 20). Gold represents registered trial design fields.

| Design field | Gold | Exact | OntSim |
|---|---|---|---|
| allocation | RANDOMIZED | RANDOMIZED | RANDOMIZED |
| masking | NONE | NONE | NONE |
| intervention model | PARALLEL | PARALLEL | PARALLEL |
| enrollment | Fewer than 50 | 50–149 | Fewer than 50 |

### 3.3 Design recommendation

Ontology-based similarity modestly improved design recommendation for previously unseen vaccine classes under leave-one-Material-out (LOMO) evaluation. We evaluated 23 ontology-connected vaccine classes, excluding all trials from one vaccine class during each training round. While Exact could still identify historical trials with similar disease, phase, and study purpose, OntSim additionally used VO relationships to identify related vaccine classes with different VO identifiers. Averaging accuracy equally across the 23 vaccine classes, OntSim achieved 0.646 compared with 0.633 for Exact (Δ = +0.013), indicating a modest improvement when identical VO identifier matches were unavailable. In a separate chronological evaluation, OntSim and Exact achieved similar field-mean accuracy (0.664 versus 0.661), while HistGBDT achieved 0.664 (**Table S3**).

The effect of ontology-based similarity on design recommendation varied across vaccine classes (**Figure 6**). OntSim improved accuracy for 11 classes, decreased accuracy for seven classes, and produced little or no change for five classes (median Δ = 0). The largest improvement was observed for autologous tumor cell vaccines (Δ = +0.125). These results suggest that ontology-based similarity can improve design recommendation when related vaccine classes share similar design characteristics, but may reduce accuracy when their historical trial designs differ.

In contrast, OntSim showed the largest decrease in accuracy for RNA vaccines (`VO_0000186`; n = 13; Δ = −0.038). Although Exact could still identify historical trials using disease, phase, and purpose, OntSim assigned higher similarity scores to some ontology-related vaccine classes with different enrollment patterns, including certain pneumococcal and DNA vaccine trials. These differences led to incorrect enrollment predictions in some cases. Complete per-class evaluation results are provided in **Table S4**.

To illustrate how ontology relationships affect individual design recommendations, we examined NCT00799110, a trial assigned to the autologous tumor cell vaccine class (`VO_0007319`), under LOMO evaluation. Both Exact and OntSim correctly predicted allocation, masking, and intervention model, but they differed in enrollment prediction. Exact predicted 50-149 participants, whereas OntSim correctly predicted fewer than 50 participants (**Table 4**). OntSim retrieved supporting GVAX vaccine trials with related but different VO identifiers (**Table S7**), illustrating how ontology-based relationships can provide additional evidence when exact VO identifier matches are unavailable.

## 4. Discussion

This study demonstrates how integrating vaccine clinical trial data into an ontology-supported SEACDM framework can support three complementary applications: trial retrieval, AE ranking, and vaccine clinical trial design recommendation. By combining the standardized data structure of SEA-CDM with semantic relationships from VO and OAE, OSEAN provides a unified resource for organizing and analyzing vaccine clinical trial information. Our results highlight the potential benefits of ontology-based integration while also showing that these benefits depend on the specific analytical task.

In Use Case 1, ontology-based query expansion enabled the retrieval of trials associated with related vaccine classes beyond exact VO identifier matching. This demonstrates the value of incorporating explicit semantic relationships into clinical trial queries. However, broader retrieval also depends on accurate ontology mappings, as incorrect assignments may introduce unrelated trials.

In Use Case 2, integrating SEA-CDM study characteristics with VO-derived vaccine families improved AE ranking compared with a frequency-based baseline. SEA-CDM provides structured trial context, including administration routes, participant characteristics, and disease indications, while VO contributes standardized vaccine classifications and biological relationships. Their combination enables contextaware AE ranking that captures information beyond overall reporting frequency, demonstrating the complementary value of structural and semantic integration. OAE further supports standardized interpretation of reported AEs. Knowledge graph embeddings also enabled the retrieval of trials sharing VO identifiers within the evaluated candidate sets.

In Use Case 3, ontology-based similarity provided modest improvements in design recommendation when identical VO identifier matches were unavailable, although performance varied across vaccine classes. However, this advantage was not observed in a separate pooled-holdout evaluation, where OntSim and Exact both achieved a trial-mean accuracy of 0.646 (**Table S5**). This suggests that the benefits of ontology-based similarity depend on the evaluation setting. This highlights that biologically related vaccine classes do not necessarily share similar trial designs. Differences in disease indications, development stages, and public health circumstances may influence enrollment strategies beyond what ontology-based similarity captures. Such contextual differences may help explain the lower performance observed for RNA vaccines, although this explanation was not directly evaluated.

Compared with previous clinical trial knowledge graph studies, our approach emphasizes the integration of structured trial information and explicit ontology relationships to support multiple downstream analyses. Chen et al. [17] developed the Clinical Trials Knowledge Graph (CTKG) to support trial similarity search and drug repurposing through graph embeddings. While their evaluation focused on embeddingbased retrieval, our integration of SEA-CDM with VO and OAE additionally supports ontology-based retrieval across related vaccine classes and context-aware AE analysis, demonstrating capabilities beyond similarity search alone. Devarakonda et al. [18] used knowledge graph embeddings and semantic inductive inference to recommend clinical trial elements, with performance evaluated primarily through textual similarity. In contrast, our approach predicts structured trial design fields and explicitly evaluates the contribution of ontology relationships when identical VO identifier matches are unavailable.

Previous work on vaccine ontology mapping and biomedical data integration established complementary foundations for our clinical trial applications. Li et al. [10] developed language model-based methods for mapping vaccine names to VO concepts, addressing the challenge of semantic standardization. Our study extends beyond concept mapping by incorporating ontology relationships into structured clinical trial data to support querying, prediction, and design recommendation. Huffman et al. [19] established SEACDM and OSEAN for integrating heterogeneous biomedical experimental data with ontology support, primarily demonstrating their application to immune-response studies. Building on this foundation, we extend the approach to clinical trial registry data and evaluate its utility across three complementary analytical applications.

The main contribution of this study is to demonstrate how the structured representation of SEA-CDM and the semantic relationships of biomedical ontologies can be jointly applied to clinical trial registry data, using vaccine clinical trials as a case study. Across three applications, this integration enabled broader ontology-based trial retrieval and improved context-aware AE ranking, while ontology-based similarity provided modest, evaluation-dependent benefits for design recommendation when exact Material matches were unavailable.

Our findings are subject to limitations related to retrospective data selection, registry-reported AE information, and the generalizability of the evaluation settings. First, our analyses relied on a retrospective AACT snapshot and keyword-based vaccine trial identification, which may have included unrelated studies or excluded relevant trials. In addition, incomplete or inaccurate VO/OAE mappings may affect ontology-based retrieval and analysis. Second, AE ranking was based on registry-reported AE terms rather than standardized incidence rates. Therefore, the predicted rankings should not be interpreted as clinical risk estimates or causal effects. Future studies could incorporate individual-level data from clinical trials and electronic health records to investigate associations between vaccine characteristics, administration routes, and AE occurrence. SEA-CDM could continue to provide a standardized structure for integrating these heterogeneous data sources, while VO and OAE could support semantic harmonization of vaccine exposures and adverse event outcomes, enabling more systematic risk factor analysis. Third, model performance and generalizability depend on the evaluation settings. Trial similarity was assessed using restricted candidate sets and shared VO identifiers as the relevance criterion, while design recommendation was evaluated on selected ontology-connected vaccine classes. The chronological evaluation used a later data snapshot and ontology release rather than a fully historical environment. Furthermore, the recommendation model has not been prospectively validated, and its outputs reflect historical trial designs rather than clinically established optimal choices.

## 5. Conclusion

In this study, we integrated vaccine clinical trial data into an ontology-supported SEA-CDM framework, combining standardized study information with semantic relationships from VO and OAE. The resulting OSEAN resource supports three complementary applications: ontology-based trial retrieval, contextaware adverse event ranking and trial similarity analysis, and clinical trial design recommendation. Our findings demonstrate that ontology relationships can improve the retrieval of related vaccine trials and support design recommendation, particularly when identical VO identifiers are unavailable, although performance varies across vaccine classes. By preserving links to the original ClinicalTrials.gov records, the framework provides traceable evidence for downstream analysis and decision support. Future work will focus on improving VO/OAE mapping coverage and accuracy, evaluating the additional contribution of OAE relationships to AE ranking, and assessing the framework through user-oriented evaluations.

## Data Availability

The clinical trial registry data analyzed in this study were obtained from the publicly available ClinicalTrials.gov database through the Aggregate Analysis of ClinicalTrials.gov (AACT) resource (snapshot dated April 12, 2026). The analysis code and supporting research materials are available on GitHub (https://github.com/zhujiadongok/osean-ctr) and Zenodo (https://zenodo.org/records/23773934). The full AACT database is not redistributed in these repositories and can be obtained from the original data provider, subject to its current access requirements. No identifiable individual-level patient data were used.

https://zenodo.org/records/23773934

https://clinicaltrials.gov/

https://aact.ctti-clinicaltrials.org/

## Funding / Data & Code Availability

### Funding

This work was supported by the National Institute of Allergy and Infectious Diseases (NIAID), National Institutes of Health (NIH), under grant U24AI171008.

### Author contributions

J. Zhu: conceptualization, methodology, software development, formal analysis, evaluation, and writing—original draft. F.-Y. Yeh: vaccine ontology mapping resources and data curation. J. Zheng: writing—review and editing. Y. He: supervision, resources, and writing—review and editing. All authors reviewed and approved the final manuscript.

### Data and code availability

The code used for data integration, analysis, and evaluation is available at https://github.com/zhujiadongok/osean-ctr (repository / package: `osean-ctr` / `ctr`). Supporting data and analysis artifacts are deposited on Zenodo (concept DOI: https://doi.org/10.5281/zenodo.23773933; version-specific DOI: https://doi.org/10.5281/zenodo.23773934). The original AACT extract is not redistributed. Details of the source snapshot and file checksums are provided in Section S6.

## References

1. Zarin DA, Tse T, Williams RJ, Califf RM, Ide NC. The ClinicalTrials.gov results database—update and key issues. N Engl J Med. 2011;364:852–860.

2. Tasneem A, Aberle L, Ananth H, et al. The database for Aggregate Analysis of ClinicalTrials.gov (AACT) and subsequent regrouping by clinical specialty. PLoS One. 2012;7:e33677.

3. Smith B, Ashburner M, Rosse C, et al. The OBO Foundry: coordinated evolution of ontologies to support biomedical data integration. Nat Biotechnol. 2007;25:1251–1255.

4. Wilkinson MD, Dumontier M, Aalbersberg IJ, et al. The FAIR Guiding Principles for scientific data management and stewardship. Sci Data. 2016;3:160018. doi:10.1038/sdata.2016.18

5. He Y, Cowell L, Diehl AD, et al. VO: Vaccine Ontology. Nature Precedings. 2009.

6. Lin Y, He Y. Ontology representation and analysis of vaccine formulation and administration and their effects on vaccine immune responses. J Biomed Semantics. 2012;3:17.

7. Zheng J, Lin AY, Huffman A, et al. VO: The Vaccine Ontology. Sci Data. 2026;13:994. doi:10.1038/s41597-026-07298-w

8. Xiang Z, Todd T, Ku KP, et al. VIOLIN: vaccine investigation and online information network. Nucleic Acids Res. 2008;36:D923–D928.

9. He Y, Racz R, Sayers S, Lin Y, Todd T, Hur J, et al. Updates on the web-based VI-OLIN vaccine database and analysis system. Nucleic Acids Res. 2014;42:D1124–D1132. doi:10.1093/nar/gkt1133

10. Li J, Li Y, Pan Y, et al. Mapping vaccine names in clinical trials to vaccine ontology using cascaded fine-tuned domain-specific language models. J Biomed Semantics. 2024;15:14. doi:10.1186/s13326-024-00318-x

11. He Y, Sarntivijai S, Lin Y, et al. OAE: The Ontology of Adverse Events. J Biomed Semantics. 2014;5:29. doi:10.1186/2041-1480-5-29

12. Pan C, Yang Q, Zhang X, Luo S, He Y, Xie J. The Ontology of Adverse Events in 2025. Sci Data. 2026;13:259. doi:10.1038/s41597-026-06584-x

13. Sarntivijai S, Xiang Z, Shedden KA, et al. Ontology-based combinatorial comparative analysis of adverse events associated with killed and live influenza vaccines. PLoS One. 2012;7:e49941.

14. Hripcsak G, Duke JD, Shah NH, et al. Observational Health Data Sciences and Informatics (OHDSI): opportunities for observational researchers. Stud Health Technol Inform. 2015;216:574–578.

15. Overhage JM, Ryan PB, Reich CG, Hartzema AG, Stang PE. Validation of a common data model for active safety surveillance research. J Am Med Inform Assoc. 2012;19:54–60. doi:10.1136/amiajnl-2011-000376

16. Tse T, Fain KM, Zarin DA. How to avoid common problems when using ClinicalTrials.gov in research: 10 issues to consider. BMJ. 2018;361:k1452.

17. Chen Z, Peng B, Ioannidis VN, et al. A knowledge graph of clinical trials (CTKG). Sci Rep. 2022;12:4724.

18. Devarakonda MV, Mohanty S, Sunkishala RR, Mallampalli N, Liu X. Clinical trial recommendations using Semantics-Based inductive inference and knowledge graph embeddings. J Biomed Inform. 2024;154:104627. doi:10.1016/j.jbi.2024.104627

19. Huffman A, Yeh F-Y, Hur J, et al. SEA CDM: Study-Experiment-Assay Common Data Model and Databases for Cross-Domain Data Integration and Analysis. Sci Data. 2026;13:238. doi:10.1038/s41597-026-06558-z

20. Miller JJ. Graph database applications and concepts with Neo4j. In: Proceedings of the Southern Association for Information Systems Conference (SAIS). 2013:24. https://aisel.aisnet.org/sais2013/24

21. Pedregosa F, Varoquaux G, Gramfort A, et al. Scikit-learn: machine learning in Python. JMLR. 2011;12:2825–2830.

22. Bordes A, Usunier N, Garcia-Duran A, Weston J, Yakhnenko O. Translating embeddings for modeling multi-relational data. In: Advances in Neural Information Processing Systems 26 (NeurIPS). 2013.

23. Vashishth S, Sanyal S, Nitin V, Talukdar P. Composition-based multi-relational graph convolutional networks. In: International Conference on Learning Representations (ICLR). 2020.

24. Resnik P. Using information content to evaluate semantic similarity in a taxonomy. In: Proceedings of the 14th International Joint Conference on Artificial Intelligence (IJCAI). 1995:448–452.

25. Pesquita C, Faria D, Falcão AO, Lord P, Couto FM. Semantic similarity in biomedical ontologies. PLoS Comput Biol. 2009;5:e1000443. doi:10.1371/journal.pcbi.1000443

26. Schlichtkrull M, Kipf TN, Bloem P, van den Berg R, Titov I, Welling M. Modeling relational data with graph convolutional networks. In: The Semantic Web (ESWC). 2018:593–607.

