## Supplementary Appendix for "Ontology-Based SEA-CDM Integration for Vaccine Clinical Trials: Query, Similarity, and Design Recommendation"

### S1. Vaccine cohort selection and feature sources

Whole-word, case-insensitive matches on AACT `studies.brief_title`, `interventions.name`, or `keywords.name` (union of NCT ids). Not used for inclusion: `intervention_type`, MeSH/`browse_*` discovery, detailed descriptions, VO ids, or LLM enrichment. Terms are taken from the VaxCT preprocessing filter (`1.filter_data.py`).

**Table S1.** Complete keyword list used for vaccine-cohort inclusion.

| Category | Terms |
|---|---|
| General | vaccine, vaccination, vaccinating, vaccinal, vaccinee, immunization, immunisation, immunize, immunise, immunizing |
| Product / sponsor tokens | pfizer, moderna, astrazeneca, johnson, novavax, biontech |
| Platform / formulation | mrna, adenovirus vector, viral vector, adjuvant |
| Disease / product phrases | covid-19 vaccine, flu vaccine, influenza vaccine, hepatitis vaccine, measles vaccine, polio vaccine |
| Pathogen / product tokens | pneumococcal, meningococcal, hpv, papillomavirus, varicella, zoster, rabies, yellow fever, typhoid, cholera, anthrax, smallpox, bcg, rotavirus, rsv, tetanus, diphtheria, pertussis, mmr, dtap, tdap, ipv, hib |

Cohort size on the analyzed snapshot: AACT `studies` = 579,828 $\rightarrow$ keyword-included vaccine NCTs = 9,147 (SEA-CDM remapped v1.4). Primary-investigational `arch_vo` assignment and remediation are described in **Section S8** and are separate from this NCT inclusion step.

**Table S2.** Feature sources used in the OSEAN analyses.

| Feature | Source file | Join | Notes |
|---|---|---|---|
| disease, phase, purpose, enrollment, start_date, design labels | `study_context_profile.csv` | NCT | Registry snapshot fields; UC3 neighbor score + gold |
| arch_vo (primary VO ID) | `disease_vo_map.csv` | NCT | product/platform/both; UC1 filter; UC2/3 VO identifier |
| vo_platform | profile platform VO + roll-up map | — | unknown if non-VO |
| sex, older, child, adult | `subject_demographics_summary.csv` | NCT | UC2 Model B context $C$ |
| route, dose | `study_intervention_profile.csv` | NCT | UC2 Model B context $C$ |
| arm_role, AE terms | Group–Occurrence arm AE index | NCT×arm | UC1 Layer A; UC2 ranking unit |
| sponsor_class | profile | — | |

Entrypoints and full paths: Section S6.

### S2. Chronological design recommendation, baselines, and validation

Main-text UC3 emphasizes Leave-one-Material-out (**Figure 6**, **Table 4**). This section holds the **chronological** past→future protocol from §2.4 / §3.3: train start <2022, test start ≥2022, so neighbors are only earlier registrations. OntSim ≈ Exact here is expected when many test VO identifiers already appear before 2022 (same-id neighbors; $s^{\text{OntSim}} = 1$). Non-ontology / text baselines formerly summarized in a main-text baselines subsection live here.

**Statistics.** Bootstrap for OntSim contrasts: 1,500 trial resamples; percentile 95% CI; seed 7. Macro-F1: sklearn `f1_score(..., average="macro", zero_division=0)` after excluding unknown gold. Prior 2022+ exploration disclosed.

**Table S3.** Chronological Setting A (n=283; `table1_lock.json`). Primary pair first; other rows are baselines.

| Method | Field-mean acc | Macro-F1 |
|---|---|---|
| OntSim kNN@20 | 0.664 | 0.332 |
| Exact-match kNN@20 | 0.661 | 0.324 |
| HistGBDT | 0.664 | 0.361 |
| Logistic regression | 0.663 | 0.355 |
| IC-sib kNN@20 (exploratory) | 0.661 | 0.324 |
| Frequency by phase | 0.633 | 0.257 |
| Title-embedding kNN@20† | 0.651 | 0.317 |
| BM25 / TF-IDF k=20† | 0.648 / 0.652 | 0.304 / 0.336 |

†Exploratory; BM25/TF-IDF k chosen on a test-period grid. Title-embedding is not a reproduction of [18].

**IC-sib (exploratory).** Motivated by information-content-based approaches to ontology semantic similarity [24,25], we used IC-weighted Jaccard similarity over subclass neighborhoods expanded by sibling classes (same direct parent; no role union). $\text{IC}(c) = -\log(|d : c \in \text{subclassClosure}(d)|/|C|)$. Gate: apply only when query VO identifier is absent from training; else exact equality. Under LOMO the gate is always open.

**Validation-tuned weights (exploratory).** On the 2019–2021 validation slice, retuning purpose/platform weights for IC-sib OntSim (purpose 1.0, platform 0.5; other weights as in §2.4) yielded chronological field-mean 0.670 and pooled trial-mean 0.638 when scored once on test/pooled. Because weights no longer match the exact-match control, this variant is exploratory only and is not used for the primary Exact vs OntSim comparison in main-text **Table 4**.

**Supervised ontology R-GCN (exploratory).** We evaluated a relational graph convolutional network (R-GCN), a model architecture introduced by Schlichtkrull et al. [26]. Entrypoint: `python -m ctr.ontology_gnn`. Chronological field-mean 0.635 with or without VO edges. LOMO class-equal: +VO 0.628 / no-VO 0.619 (Δ +0.009, seed 7). Main-text Leave-one-Material-out contrast remains OntSim vs Exact (**Figure 6**, **Table 4**).

### S3. Resource counts, OAE coverage, and structural audit

Studies 9,147; experiments 18,294; subjects 9,147; interventions 20,001; occurrence 617,510; results 1,525,757. CTR n=1,418.

**OAE AE grounding (shared OSEAN graph).** Distinct registry AE terms with an OAE id: 5,040 / 14,190. Of the locked UC2 ranking-label set, 78 / 80 terms are OAE-grounded. Graph edges: `AE_TERM`–`ae_term-grounded_in_oae`–`OAE` plus `oae-subClassOf` ancestors (export: `osean_kg/osean_oae.tsv`; term map: `osean_kg/ckpts/sea_ml_v14/ae_oae_map.csv`). OAE is used for inspectable AE labels in UC1/UC2; it is not a design-field ontology for UC3.

**SEA-CDM structural audit.** See `optimize/seacdm_linkage/` and `v1.4/OFFICIAL_V14_*`. Spine foreign keys are 100% row-resolved in the remapped vaccine package.

### S4. LOMO eligibility, per-class results, and pooled holdout

**Eligible vaccine classes.** Leave-one-Material-out evaluation included 23 vaccine classes with 12–80 trials each and at least one ontology relationship to another vaccine class in the cohort. After primary-investigational mapping remediation this yields the 23 classes below (labels from `vo.owl` via `vo_eligible_labels.py`). The eligible classes, sample sizes, and per-class evaluation results are reported in the table.

Main-text LOMO reports class-equal OntSim 0.646 vs Exact 0.633 ($\Delta$ +0.013; **Figure 6**). Trial-equal on the same 23 vaccine classes: OntSim 0.648, Exact 0.640 ($\Delta$ +0.008); frequency 0.645; HistGBDT 0.659; IC-sib 0.656 (`after_vo_fix_eval.json`). Bootstrap settings for LOMO contrasts match Section S2. Per-class trial-mean accuracy:

**Table S4.** Per-class trial-mean accuracy under leave-one-Material-out evaluation.

| VO id | Label | n | Exact | OntSim | IC-sib | Freq. | HistGBDT |
|---|---|---|---|---|---|---|---|
| VO_0000534 | subunit vaccine | 80 | 0.647 | 0.641 | 0.619 | 0.641 | 0.644 |
| VO_0000561 | toxoid vaccine | 77 | 0.689 | 0.686 | 0.686 | 0.676 | 0.696 |
| VO_0000032 | DNA vaccine | 75 | 0.653 | 0.677 | 0.680 | 0.630 | 0.663 |
| VO_0004673 | quadrivalent influenza vaccine | 67 | 0.649 | 0.646 | 0.646 | 0.657 | 0.731 |
| VO_0000367 | live attenuated vaccine | 66 | 0.620 | 0.639 | 0.663 | 0.631 | 0.645 |
| VO_0001178 | live attenuated influenza vaccine | 60 | 0.582 | 0.590 | 0.604 | 0.637 | 0.604 |
| VO_0003150 | Hepatitis B surface antigen vaccine | 58 | 0.621 | 0.638 | 0.655 | 0.621 | 0.677 |
| VO_0000484 | recombinant vector vaccine | 57 | 0.697 | 0.697 | 0.719 | 0.678 | 0.711 |
| VO_0000642 | influenza virus vaccine | 56 | 0.641 | 0.637 | 0.637 | 0.624 | 0.624 |
| VO_0000771 | BCG vaccine | 51 | 0.675 | 0.675 | 0.685 | 0.673 | 0.649 |
| VO_0000761 | malaria vaccine | 31 | 0.548 | 0.589 | 0.637 | 0.589 | 0.653 |
| VO_0006034 | meningococcal vaccine (unknown formulation/serogroups) | 31 | 0.629 | 0.621 | 0.621 | 0.645 | 0.653 |
| VO_0000047 | Fluzone | 28 | 0.580 | 0.598 | 0.607 | 0.580 | 0.625 |
| VO_0000577 | peptide vaccine | 18 | 0.764 | 0.736 | 0.764 | 0.750 | 0.778 |
| VO_0003075 | Gardasil 9 vaccine | 18 | 0.625 | 0.639 | 0.653 | 0.667 | 0.611 |

| VO id *(continued)* | Label | n | Exact | OntSim | IC-sib | Freq. | HistGBDT |
|---|---|---|---|---|---|---|---|
| VO_0007062 | Quadrivalent HPV (6/11/16/18) recombinant vaccine | 17 | 0.647 | 0.647 | 0.632 | 0.750 | 0.603 |
| VO_0004246 | Anthrax vaccine adsorbed with squalene adjuvant | 16 | 0.594 | 0.609 | 0.625 | 0.578 | 0.609 |
| VO_0007319 | autologous tumor cell vaccine | 16 | 0.651 | 0.776 | 0.760 | 0.667 | 0.682 |
| VO_0005158 | Oxford-AstraZeneca COVID-19 vaccine | 15 | 0.583 | 0.600 | 0.617 | 0.567 | 0.567 |
| VO_0007063 | Recombinant HPV nonavalent vaccine | 14 | 0.625 | 0.625 | 0.607 | 0.625 | 0.661 |
| VO_0000186 | RNA vaccine | 13 | 0.673 | 0.635 | 0.615 | 0.615 | 0.558 |
| VO_0003464 | Hepatitis A vaccine, inactivated | 12 | 0.625 | 0.708 | 0.667 | 0.646 | 0.688 |
| VO_0003834 | poliovirus vaccine inactivated | 12 | 0.542 | 0.542 | 0.646 | 0.688 | 0.646 |

**Table S5.** Pooled holdout of the same 23 classes (train 530 / test 888)—supplement only; methods converge and are not used as a primary ontology claim.

| Method | Trial-mean | Field-mean |
|---|---|---|
| Exact-match kNN@20 | 0.646 | 0.644 |
| OntSim kNN@20 | 0.646 | 0.643 |
| IC-sib kNN@20 | 0.644 | 0.642 |
| Frequency by phase | 0.632 | 0.630 |
| HistGBDT | 0.615 | 0.613 |

### S5. Use case 2 similarity ablations

Main-text **Table 2** reports locked AE recall@10 (Model A 0.317 → Model B 0.383; `ckpts/sea_ml_v14/metrics.json`) and SEA-CDM+VO trial-similarity Hits/MRR. The Comirnaty-style Model A → Model B re-ranking example is main-text **Table 3**. OAE ids label predicted/gold AE strings for inspection (78/80 ranking labels); they are not Model B input features, and no OAE-aware AE-ranking ablation is reported here (reserved for future work).

**Trial similarity VO-only.** Same split as **Table 2** (same VO identifier; candidates in disease box; n=541), but the graph keeps VO `subClassOf/has_role` edges and drops SEA-CDM instance edges. TransE Hits@1/10/MRR 0.699/0.972/0.800; CompGCN 0.479/0.891/0.621. Main text headlines the **full** SEA-CDM+VO graph only (OAE edges are not required for these locked Hits/MRR numbers).

**AE ranking alternate split.** An older NCT-grouped protocol (n_eval=483) is not pooled with the primary study-grouped split used for Table 2 / Table 3.

### S6. Reproducibility file inventory

Public code: https://github.com/zhujiadongok/osean-ctr (package ctr). Supporting data: https://doi.org/10.5281/zenodo.23773933

AACT snapshot. Pipe-delimited ClinicalTrials.gov / AACT extract dated 2026-04-12.

- Zip path: AACT Database/6wvqgoof4dut7eaxn610uzfajvnt.zip (filesystem mtime 2026-04-12).
- studies.txt row version / update timestamps 2026-04-12 04:05:04.982706.
- Local folder id 6wvqgoof4dut7eaxn610uzfajvnt; studies.txt size 390,802,838 bytes.
- SHA-256: 82870fc4a17cc1eb 75529c603866db82 207ff7366ab49569 f4779f89ffaba7f8.

VO OWL. Release 2026-06-12 under:

SEA-CDM AACT vaccine remapped/research_factors/vo_mapping/FOR_ADVISOR/vo.owl.

OAE graph export / AE term map (coverage in S3): osean_kg/osean_oae.tsv, osean_kg/ckpts/sea_ml_v14/ae_oae_map.csv (import helper: osean_neo4j/import_oae_to_neo4j.py).

- Table 1 lock (chrono): optimize/table1_lock.json (python -m ctr.eval_table1_lock)
- Post-remediation neighbors / LOMO: optimize/mapping_audit/after_vo_fix_eval.json
- LOMO mechanism example (NCT00799110): optimize/mapping_audit/mechanism_example_NCT_neighbors.json
- Three use-case structure examples: optimize/mapping_audit/use_case_structure_examples.json
- Trial similarity scores: ckpts/sea_ml_v14/seacdm_similarity.txt (+ seacdm_similarity_examples.csv)
- AE ranking bundle: ckpts/sea_ml_v14/sea_ml_phase1.joblib (+ metrics.json)
- Mapping audit: optimize/mapping_audit/ (validate_vo_mapping must pass)
- Entrypoints: eval_table1_lock; eval_after_vo_fix; export_mechanism_example; validate_vo_mapping; apply_vo_corrections_upstream
- Demo / prototype: osean_neo4j/app_xy_isg.py (Figure S1)

### S7. Prototype Design interface

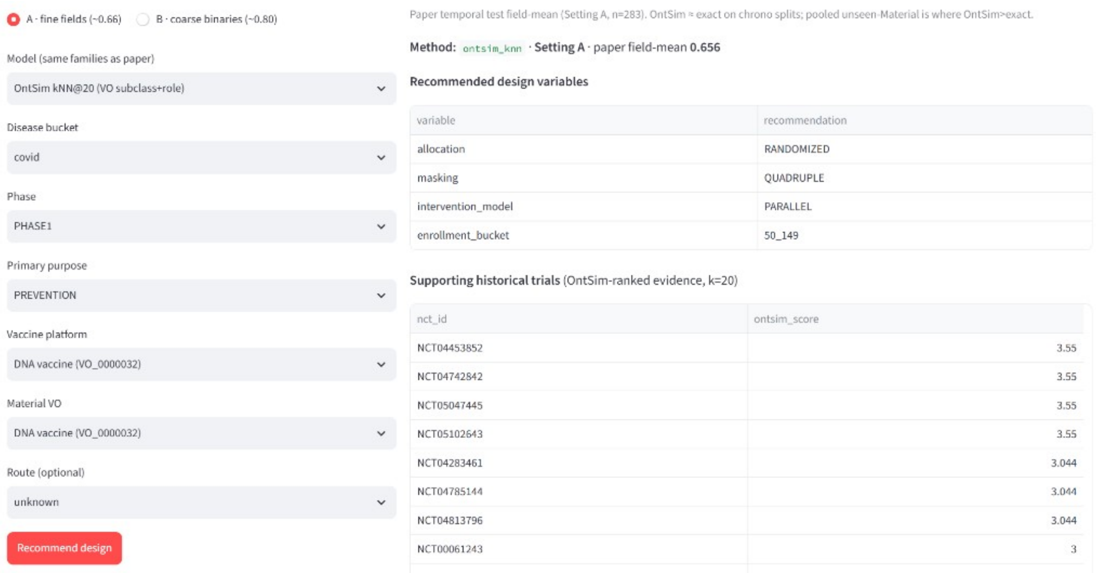


**Figure S1**. Prototype Design view: query inputs, predicted design fields, and OntSim-ranked supporting NCT ids. Decision support only.

### S8. Mapping audit and primary-investigational corrections

**Primary-investigational mapping (locked).** `arch_vo` = primary investigational vaccine only; `vo_platform` = that vaccine's technology platform.

Rubric. `correct` / `incorrect` / `ambiguous` for Material and platform separately; `unknown` platform = unlabeled.

Audit sample (`mapping_audit_sample_reviewed.csv`, n=80). Disposition by reviewer tag (not interchangeable with expert correctness):

- `advisor_review`: 19 (human advisor seed)
- `ctgov_final22`: 22 (CT.gov-checked final-22 set)
- `auto_remediated_verified`: 39 (auto-remediation + programmatic verification)

Judgment counts describe the mappings at the time of review, before application of the final corrections. Material: correct 60 / incorrect 17 / ambiguous 2 / blank 1. Platform: correct 59 / incorrect 12 / ambiguous 4 / blank 5. These support "80 rows have disposition records," not "80/80 expert-verified correct." `validate_vo_mapping` PASS confirms regression checks (overlay presence, intentional unknowns, rule integrity)—not independent semantic accuracy for every CTR row.

Expanded remediation (620 NCTs, ~44% of CTR). Targets in `vo_corrections_primary.csv` applied upstream (`disease_vo_map` + `study_context_profile`) and as an idempotent CTR overlay. Hard checks: `python -m ctr.validate_vo_mapping` (must pass). Rebuild: `build_vo_corrections_bulk` → `apply_vo_corrections_upstream` → `validate_vo_mapping` → `build_table` → `eval_after_vo_fix` / `eval_table1_lock`.

Cohort mapping how / platform (CTR n=1,418). Pre-remediation snapshot: how platform 801 / product 320 / both 297; `vo_platform=unknown` 241. Post-remediation (analysis table): how platform 539 / product 101 / both 778; unknown 24.

Post-remediation neighbor metrics. `after_vo_fix_eval.json` + `table1_lock.json`. Chrono exact=IC-sib 0.661 / role 0.664 / HistGBDT 0.664; pooled ~0.645 (n=888, 23 classes); LOMO class-equal exact 0.633 / OntSim 0.646 / IC-sib 0.654 / HistGBDT 0.651 / frequency 0.645.

### S9. Query Layer A—exact subunit AE ranking (OAE-grounded)

When the VO identifier is fixed (subunit `VO_0000534`; 166 mapped NCTs; 39 vaccine arms with AE reports), Layer A returns NCT-traceable AE counts with OAE labels (not predicted rates):

**Table S6.** Exact VO identifier query for subunit vaccine AEs with OAE labels.

| rank | ae_term | trial_count | OAE id | OAE label |
|---|---|---|---|---|
| 1 | Headache | 19 | OAE_0000377 | headache AE |
| 2 | Vomiting | 17 | OAE_0000601 | vomiting AE |
| 3 | Myalgia | 17 | OAE_0000375 | myalgia AE |
| 4 | Cough | 17 | OAE_0000398 | cough AE |
| 5 | Fatigue | 16 | OAE_0000034 | fatigue AE |
| 6 | Nausea | 15 | OAE_0000600 | nausea AE |
| 7 | Pyrexia | 14 | OAE_0000361 | fever AE |
| 8 | Diarrhoea | 13 | OAE_0000616 | diarrhea AE |

### S10. OntSim supporting neighbors for `NCT00799110` (LOMO)

Top OntSim neighbors for the Table 4 query trial `NCT00799110` when its autologous tumor cell vaccine class (`VO_0007319`) is held out; neighbor scores are from `mechanism_example_NCT_neighbors.json` (GVAX neighbors are ontology-related supports, not the query vaccine class).

**Table S7.** Representative supporting trials retrieved by OntSim.

| supporting NCT | Neighbor VO class | OntSim | same id? | $S(q, j)$ |
|---|---|---|---|---|
| NCT00569387 | VO_0007610 GVAX pancreatic | 0.438 | no | 3.719 |
| NCT00614601 | VO_0007610 GVAX pancreatic | 0.438 | no | 3.719 |
| NCT00727441 | VO_0007610 GVAX pancreatic | 0.438 | no | 3.719 |
| NCT00116467 | VO_0007467 GVAX cancer | 0.368 | no | 3.684 |
| NCT00771017 | VO_0007467 GVAX cancer | 0.368 | no | 3.684 |

### S11. Full RNA-platform expand delta (VO_0000186; +25)

Main-text **Table 1** shows six representatives. Below is the complete expand-minus-exact set (16 → 41): VO_0005198 COVID-19 RNA roll-up (n=20) and VO_0007132 RNA-related descendant (n=5).

**Table S8.** Complete RNA-platform ontology-expansion delta (+25 trials).

| Trial | Product | Stored VO ID | Why missed by exact id |
|---|---|---|---|
| NCT04566276 | ChulaCov19 | VO_0005198 | COVID-19 RNA roll-up |
| NCT04780659 | Comirnaty | VO_0005198 | COVID-19 RNA roll-up |
| NCT04826770 | mRNA-1273.351 | VO_0005198 | COVID-19 RNA roll-up |
| NCT04834869 | mRNA-1273.351 | VO_0005198 | COVID-19 RNA roll-up |
| NCT04860297 | mRNA-1273.351 | VO_0005198 | COVID-19 RNA roll-up |
| NCT04930770 | mRNA-1273.351 | VO_0005198 | COVID-19 RNA roll-up |
| NCT04934111 | LNP-nCOV saRNA-02 | VO_0005198 | COVID-19 RNA roll-up |
| NCT05012943 | ARCT-154 | VO_0005198 | COVID-19 RNA roll-up |
| NCT05054218 | mRNA-1273.351 | VO_0005198 | COVID-19 RNA roll-up |
| NCT05145348 | Comirnaty | VO_0005198 | COVID-19 RNA roll-up |
| NCT05283902 | Comirnaty | VO_0005198 | COVID-19 RNA roll-up |
| NCT05305573 | Comirnaty | VO_0005198 | COVID-19 RNA roll-up |
| NCT05329064 | Comirnaty | VO_0005198 | COVID-19 RNA roll-up |
| NCT05370040 | AAHI-SC2 | VO_0005198 | COVID-19 RNA roll-up |
| NCT05405283 | Comirnaty | VO_0005198 | COVID-19 RNA roll-up |
| NCT05435027 | GRT-R912 | VO_0005198 | COVID-19 RNA roll-up |
| NCT05542693 | Comirnaty | VO_0005198 | COVID-19 RNA roll-up |
| NCT05605470 | ChulaCov19 | VO_0005198 | COVID-19 RNA roll-up |
| NCT05697705 | Bivalent Omicron BA.1–adapted BNT162b2 | VO_0005198 | COVID-19 RNA roll-up |
| NCT06279871 | 2008–2009 trivalent influenza vaccine | VO_0005198 | COVID-19 RNA roll-up |
| NCT02074046 | mRNA-loaded autologous DC vaccine | VO_0007132 | RNA-related descendant |
| NCT02084823 | mRNA-loaded autologous DC vaccine | VO_0007132 | RNA-related descendant |
| NCT02089919 | mRNA-loaded autologous DC vaccine | VO_0007132 | RNA-related descendant |
| NCT02115958 | mRNA-loaded autologous DC vaccine | VO_0007132 | RNA-related descendant |
| NCT02176746 | mRNA-loaded autologous DC vaccine | VO_0007132 | RNA-related descendant |
